# Multi-Ancestry Rare and Low-Frequency Variant Analysis Reveals Novel and Convergent Biological Pathways for Cannabis Use Disorder

**DOI:** 10.1101/2025.03.16.25324078

**Authors:** Qian Peng, Xiwen Zhang, Kirk C. Wilhelmsen, Cindy L. Ehlers

## Abstract

Large disparities in Cannabis Use Disorder (CUD) prevalence exist across U.S ethnic groups, yet genetic research has disproportionately focused on individuals of European descent. This study investigated the genetic architecture of CUD severity across three populations—American Indians, Mexican Americans, and European Americans—focusing on rare and low-frequency variants (minor allele frequency < 5%). CUD severity was measured using a validated composite of 20 cannabis-related life events, enhancing phenotypic resolution and power. In American Indians, ten genome-wide significant common variants were identified. Analyses of rare and low-frequency functional variants revealed novel, convergent biological pathways across ancestries. Notably, the arylsulfatases activation pathway was significantly associated with CUD severity in Mexican Americans and supported in American Indians, implicating sulfated lipids such as sphingolipids in CUD neurobiology. Related, in European Americans, the heparan sulfate degradation pathway—central to synaptic remodeling and extracellular matrix regulation—showed significant negative association with CUD severity. The integrin beta-1 cell surface interaction pathway, critical for neuronal adhesion and signaling, was uniquely enriched in Mexican Americans. Immune-related pathways, including shared involvement of a p38-gamma/delta MAPK signaling cascade, were supported across multiple cohorts. These findings highlight both ancestry-specific and shared mechanisms contributing to CUD severity, emphasizing the biological complexity of addiction. Importantly, these pathways represent potential therapeutic targets, suggesting that modulation of sulfated lipid metabolism and extracellular matrix remodeling may offer novel strategies for intervention. By integrating multi-ethnic cohorts and rare variant analyses, this study identifies underappreciated genetic contributors to CUD and underscores the importance of population-informed addiction genetics research.

## INTRODUCTION

Cannabis stands as the most frequently utilized psychotropic substance in the United States (12-month use prevalence of 16%), following alcohol. There is a noticeable increase in its usage, particularly among the youth [SAMHSA, 2018]. As more states legalize recreational cannabis use and the perception of its harm diminishes, there is a potential for a growing population of cannabis users, leading to higher rates of adverse health consequences [Volkow et al., 2014]. Approximately one-fifth of lifetime users meet criteria for DSM-5 cannabis use disorder (CUD), with nearly a quarter classified as severe [Hasin, 2018]. Consistent findings from studies on twins and families indicate a genetic influence on the development of cannabis use and use disorders [Agrawal and Lynskey, 2006], with problematic use being heritable at 50-60% [Verweij et al., 2010]. Epidemiology studies have also found considerable variation in the prevalence of CUD across different ethnic groups [Hasin, 2018]. For instance, American Indians have the highest rates (11.5%) of CUD in the U.S., especially severe cases. The proportion of CUD variability attributable to common genomic variants (SNP-based heritability) varies across ethnic groups as well, ranging from 6.7% in individuals of European descent to 18% in those of Hispanic and Latino Americans [Levey et al., 2023]. Thus, it seems that different ancestral groups may have distinct genetic risk or protective factors for CUD. The distinct genetic factors may underly different or similar biological pathways that lead to CUD.

Genome-wide association studies (GWAS) have become more effective in identifying genetic factors for CUD as sample sizes grow larger [Demontis et al., 2019; Hatoum et al., 2022; Johnson et al., 2020; Levey et al., 2023]. However, most of the findings still primarily come from European populations due to their much larger sample sizes. For instance, the largest GWAS meta-analysis to-date, which included over a million individuals, identified 22 significant loci for CUD in European descents and only one or two in other populations [Levey et al., 2023]. While GWAS mainly focus on common genomic variants, recent research highlights the importance of rare variants, particularly those in low linkage disequilibrium (LD) with neighboring variants, in contributing to complex traits and diseases. This effect is greater for protein-altering variants, which are consistent with negative selection pressure [Wainschtein et al., 2022]. Rare variants are often recent mutations and are typically weakly correlated (low LD) with other variants. In fact, most human variants are rare, as illustrated by a TOPMed study of over 53K individuals, where 97% of over 400 million detected variants had minor allele frequencies (MAF) of less than 1% [Taliun et al., 2021]. Functional variants that alter gene function are also uncommon [Kryukov et al., 2007].

In this study, we analyzed three populations, European Americans (EA), and two understudied groups: American Indians (AI) and Mexican Americans (MA). The lifetime prevalence of CUD is highest in the AI population (39%), followed by MA (27%) and EA (21%) (Figure 1). While we examined common variants, our primary focus was on rare and low-frequency variants (MAF < 5%). We hypothesized that these populations have both unique and shared genetic factors for CUD, with rare variants potentially differing across ancestries but still contribute to the same genes or similar underlying pathways. To test this, we conducted both gene-based and pathway-based analyses on these variants. Low-frequency variants were included in the analyses because, in relatively isolated populations like American Indians, some rare variants that are rare in the general population may be more common due to selection, genetic drift, or inbreeding effects. This can enhance the power of association studies [Uffelmann et al., 2021]. We used a CUD severity phenotype to quantify the progression and severity of the disorder [Ehlers et al., 2015; Peng et al., 2020]. While the severity phenotype is highly correlated with DSM-5 CUD diagnosis, it offers greater statistical power for association analyses as a quantitative trait.

**Figure 1.**
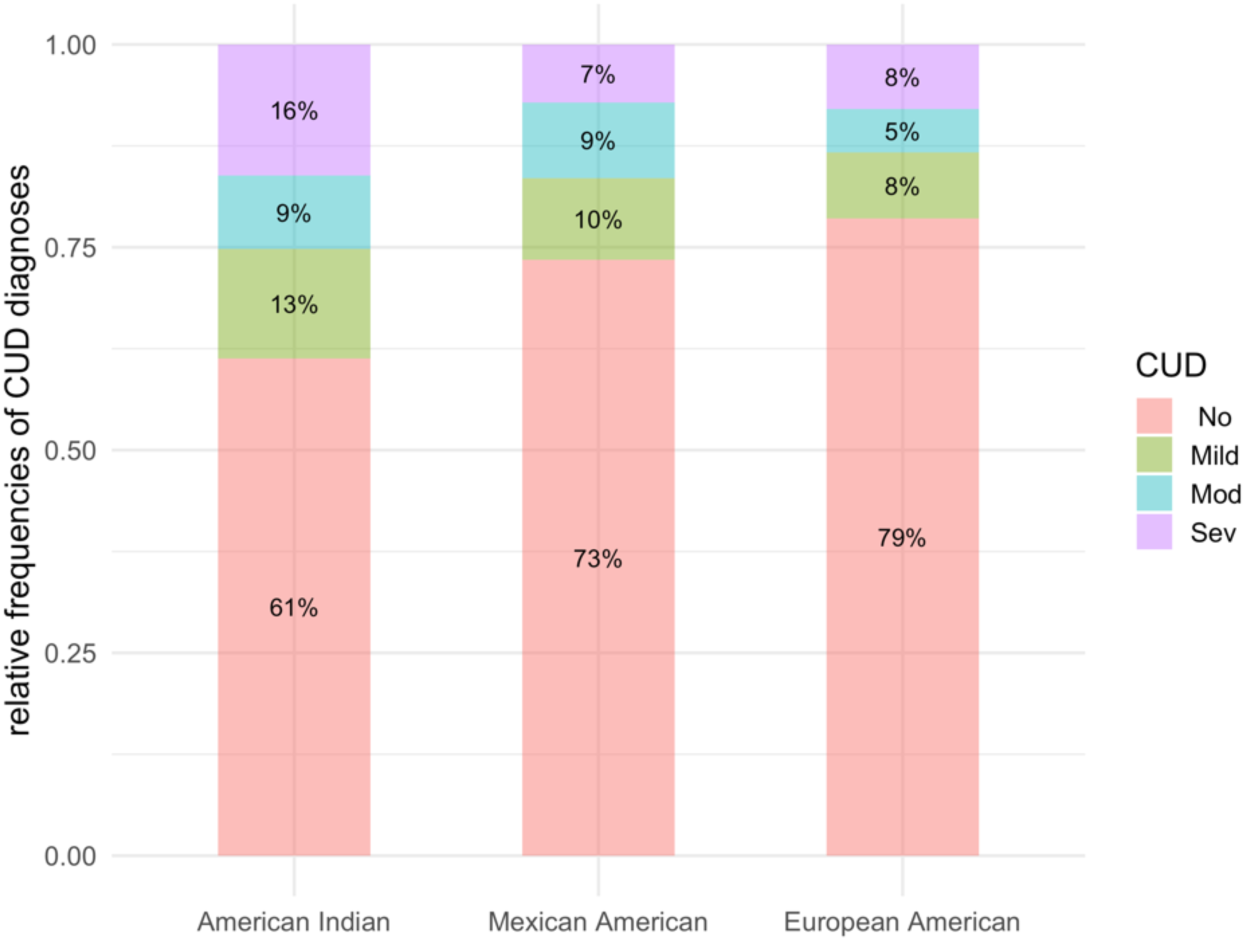
Prevalence of lifetime CUD diagnoses across American Indian, Mexican American, and European American cohorts.

## MATERIALS AND METHODS

### Participants

Three independent populations were investigated: 742 Native Americans from extended pedigrees from an American Indian cohort (AI), 547 Mexican Americans from a primarily second-generation young adult Mexican American cohort (MA), and 1711 primarily European American (EA) participants from the San Francisco Family Alcohol Study. We refer to the first cohort as AI, the second as MA, and the third as EA. Their demographics are presented in Table S1. The population characteristics and the recruitment procedures of the three cohorts have been respectively described [Ehlers et al., 2010a; Ehlers et al., 2004a; Ehlers and Gizer, 2013; Ehlers et al., 2011; Ehlers et al., 2004b; Gizer et al., 2011; Seaton et al., 2004; Vieten et al., 2004]. Details are given in Supplemental Methods. The protocol of the study of the American Indian cohort was approved by the Scripps Research Institute Institutional Review Board (TSRI-IRB) and presented to the Indian Health Council, a tribal review group overseeing health issues for the reservations where recruitments took place. TSRI-IRB also approved the protocol of the study of the MA cohort. The protocol for collection of participants in the EA cohort was approved by the University of California San Francisco (UCSF) Committee for the Protection of the Rights of Human Subject while the recruitment took place. Subsequently, the University of North Carolina, at Chapel Hill IRB approved the data analysis plan. Written informed consent was obtained from each participant after study procedures had been fully explained. Participants were compensated for their time spent in the study.

### Phenotypes

All participants were evaluated using Semi-Structured Assessment for the Genetics of Alcoholism (SSAGA) [Bucholz et al., 1994; Hesselbrock et al., 1999], from which the phenotypes were derived. We adapted measures of the clinical course of alcohol use disorder, as initially described by Schuckit and colleagues [Schuckit et al., 1993], to quantitate life events during the clinical course of other substance use disorders. These measures were based on the relative order of major “substance-related life events” that occurred during the clinical course. Previous research has shown that these life events are highly similar and consistent across various subgroups and populations, although there may be differences in the age of onset and the endorsement rates of individual events [Ehlers et al., 2010b; Ehlers et al., 2015; Ehlers et al., 2004b; Malcolm et al., 2006; Montane-Jaime et al., 2008; Schuckit et al., 1995; Schuckit et al., 2002; Scott et al., 2008; Venner and Miller, 2001]. Table S2 outlines 20 cannabis-related life events that occur during the clinical course of CUD. The sequence of these events within each population cohort was determined by the mean age at which they occurred, with the first event representing the earliest occurrence and the 20^th^ event representing the latest in the individual’s lifetime. The exact order of events thus may vary between different population cohorts. Next, we developed a metric to quantify the severity of CUD progression based on the sequential occurrence of these events. Each CUD-related life event was assigned a severity weight, as outlined in Table S2: events 1-7 received a weight 1, events 8-14 received a weight 2, and 15-20 received a weight 3. This metric assigns higher weights to the more severe events that occur later in the clinical course of CUD, which are more closely associated with severe use disorder [Ehlers et al., 2015; Peng et al., 2019]. The CUD severity phenotype was defined as the total sum of the severity weights of the life events that occurred for each individual [Peng et al., 2019]. The distributions of this phenotype for each cohort and their relationships to the DSM-5 CUD diagnoses are shown in Figure S1. The Spearman’s rank correlation (ρ) between the CUD severity phenotype and the CUD diagnosis (no, mild, moderate, and severe) is 0.90 for AI, 0.93 for MA, and 0.89 for EA. For subsequent analysis, we applied winsorization to the CUD severity at the 5% level at each tail.

### Sequencing and association analysis

The AI and EA participants had low-coverage whole genome sequencing on blood-derived DNAs with the same pipeline [Bizon et al., 2014]. The MA participants had exome genotyping imputed using the TOPMed reference panel. Details are given in SI methods.

A mixed linear model-based association analysis as implemented in GCTA-MLMA [Yang et al., 2011; Yang et al., 2014] was used to conduct the whole genome (or exome) association analysis on variants with minor allele frequency (MAF) greater than 1% for each cohort, to control for population structures and familial relatedness. The association for each variant was conditioned on a genetic relationship matrix (GRM) of the cohort derived from genotypes, thus capturing a wide range of sample structures [Yang et al., 2011].. We further included sex, age, age-squared as covariates.

### Gene-based rare and low-frequency variant analysis

Rare and low-frequency (MAF < 5%) variants were analyzed using two methods, combined multivariate and collapsing (CMC) method [Li and Leal, 2008] and a non-burden sequence kernel association test (SKAT) [Wu et al., 2011], within a mixed linear model as implemented in EPACTS [Kang et al., 2010]. For each gene, we formed two types of groups. One group considered all variants on exons, 5’ and 3’ UTRs, upstream and downstream of the gene (denoted as *ExonReg*). The other group included only the nonsynonymous variants and the splicing-site variants of the gene (denoted as *Nonsyn*). Intergenic and intronic variants were excluded in this study. For each group type, a gene was excluded if fewer than three markers were found, or if less than 0.5% of the samples had any such markers on the gene. The gene-based tests were performed on the CUD severity phenotype. False discovery rates (FDR) controlled by Benjamini–Hochberg procedure (Benjamini and Hochberg, 1995) were used to set significant thresholds (FDR<0.05) from the test statistics of the association tests. We combined four groups together (two testing methods on two gene variant groups) for the correction and report the FDR-adjusted *p* values (Yekutieli and Benjamini, 1999). The correction is conservative since the *Nonsyn* variants on a gene are a subset of the *ExonReg* variants thus the two variant groups are correlated.

### Pathway-based rare and low-frequency variant analysis

We downloaded a collection of canonical pathways from the Molecular Signatures Database (MSigDB) version 7.5.1 [Liberzon et al., 2015; Subramanian et al., 2005]. The collection is comprised of pathways from multiple curated databases including BioCarta [Nishimura, 2001], KEGG [Kanehisa et al., 2016], PID [Schaefer et al., 2009], Reactome [Gillespie et al., 2022], and WikiPathways [Kutmon et al., 2016]. Each pathway is made of a gene set. The gene sets are canonical representations of a biological process compiled by domain experts. There are total of 2981 canonical pathway gene sets in this release. Similar to the gene-based test, for each pathway gene set, we formed two variant groups: *ExonReg* and *Nonsyn*, using rare and low-frequency variants. Since pathways may contain large numbers of genes, to limit the number of variants included in each set, we limited the minimum MAF to at least 0.1%. We performed CMC and SKAT tests on each variant group from each pathway. FDR-adjusted *p* values are reported combining the four test groups for each cohort. One performance issue we encountered was that several pathways, containing a large number of genes and thus a very high number of variants, failed to converge and were therefore excluded. However, these pathways are typically higher-level pathways within the pathway hierarchy.

### Networks of pathways

After ranking pathways by how significant they were associated with CUD severity in each cohort, we examined whether top-ranked pathways were distinct or shared across the population cohorts. From each cohort, we selected the top 10 pathways linked to CUD severity from each of the four test groups. To assess the similarity between any two pathways, we used the Jaccard coefficient [Jaccard, 1901], which calculated the fraction of genes common to both pathways relative to the total number of genes in at least one of the two pathways. The Jaccard coefficient ranges from 0 (no overlap between the two pathways) to 100% (identical pathways). The similarity measure was then applied to construct pathway networks across cohorts. By setting a similarity threshold, we could identify whether network clusters of overlapping pathways emerge from different populations related to CUD severity. For this analysis, we set the Jaccard coefficient threshold to 10%.

For each such network cluster, we tested tissue specificity using differentially expressed genes within the cluster with FUMA version 1.5.2 [Watanabe et al., 2017]. The test compared gene expression levels in a specific tissue sample to all other samples, identifying genes with significantly upregulation or downregulation. The analysis used tissue-specific transcriptome data from 54 tissue types in GTEx v8 [GTEx Consortium, 2015], with samples presumed to be from healthy donors and without associated phenotypes. The goal was to assess whether the genes with the network cluster had tissue specificity that could link them to CUD. Only the most significant upregulation or downregulation was reported for genes with both.

## RESULTS

Although our GWAS analysis was primarily exploratory (see Figures S2 and S3 for Manhattan and q-q plots) due to moderate sample sizes, we identified 10 significant loci associated with CUD severity in the American Indian cohort (Table S3). The top variant, rs200530573 (*p*=5E-11, MAF=5.2%), lies downstream of the gene *NDUFA12* (NADH-ubiquinone oxidoreductase subunit A12) on chromosome 12 (Figure S4). Another significant locus is located on chromosome 19, with the leading variant rs79550841 (*p*=3.9E-8, MAF=1.5%). This locus is situated downstream of *ZNF579* and upstream of *SBK3* (Figure S4).

Gene-based analysis of rare and low-frequency variants identified *ARSA*, which encodes the enzyme arylsulfatase A, as being significantly associated with CUD severity in the MA cohort (*ExonReg* SKAT; *p*=1.62E-6, FDR=0.04) (see Figure 2, Table S4). However, no gene remained genome-wide significant after FDR correction in the AI and EA cohorts. In the European American cohort, rare and low-frequency variants in the glycosaminoglycan heparan sulfate biosynthesis pathway were significantly associated with CUD severity after multiple comparison correction (*ExonReg* CMC; *p*=1.62E-7, FDR=0.002) (see Figure 3, Table S5). In the Mexican American cohort, three pathways remained significant (Figure 3, Table S5). These included pathways related to the activation of arylsulfatases (*ExonReg* SKAT; *p*=1.57E-6, FDR=0.018), beta 1 integrin cell surface interactions (*nonsyn* CMC; *p*=8.81E-6, FDR=0.04), and an inflammatory condition, inclusion-body myositis (*ExonReg* CMC; *p*=1.10E-5, FDR=0.04). A pathway related to protein import into peroxisomal membrane was suggestively associated with CUD severity in the American Indian cohort (*ExonReg* CMC; *p*=3.09E-5). Additionally, two pathways related to the activation of arylsulfatase (*nonsyn* CMC) were among the top pathways detected in the American Indians and were nominally associated with CUD severity (*p*=4.56E-4, 2.96E-3).

**Figure 2.**
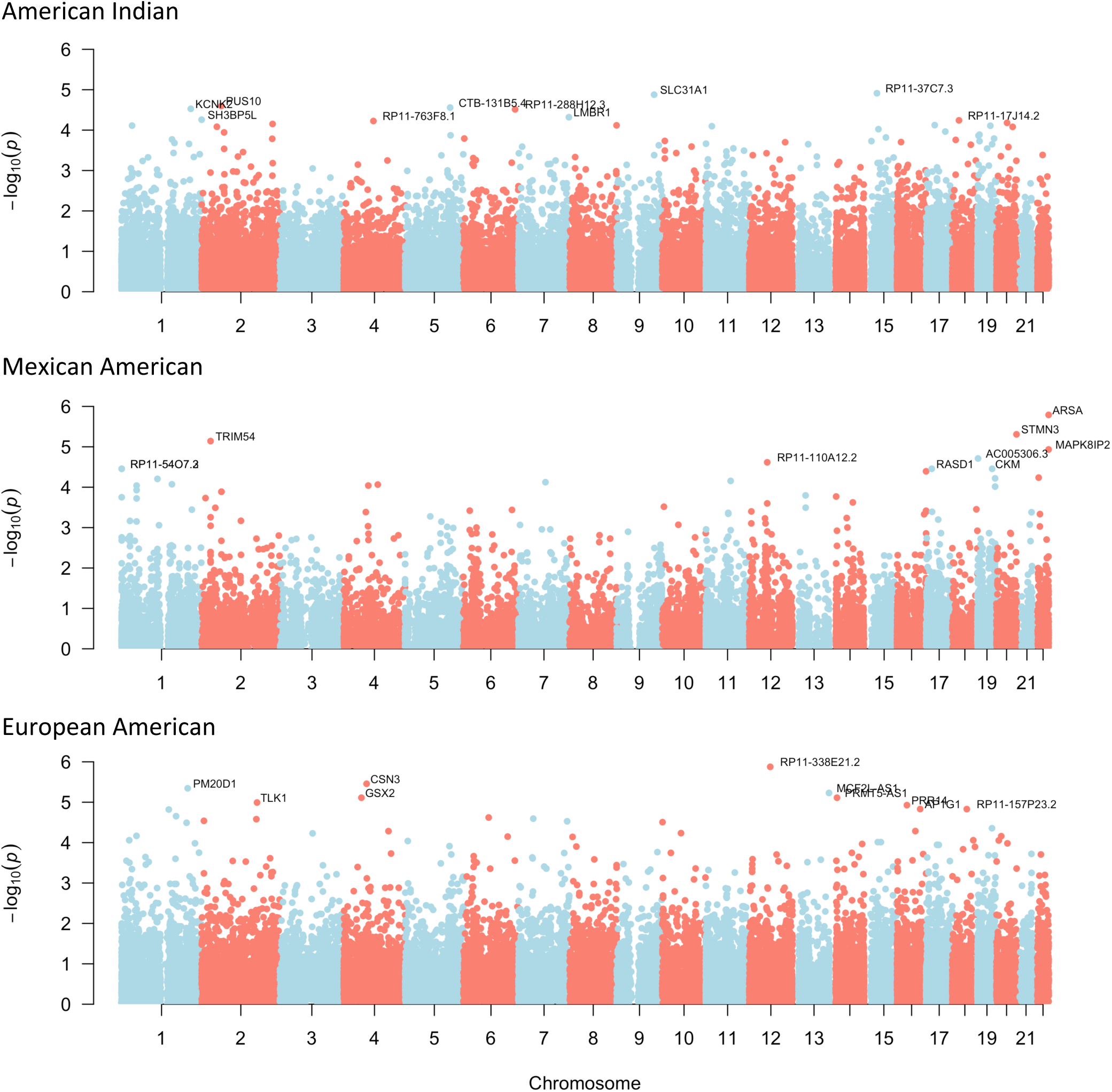
Gene-based Manhattan plots of rare and low-frequency variant associations with CUD severity across three population cohorts. For each gene, the lowest *p*-value among the four combinations of variant groups (ExonReg and Nonsyn) and association tests (CMC and SKAT) is shown. Full results are provided Table S3. After false discovery rate (FDR) correction, no genome-wide significant gene associations were observed in the AI and EA cohorts, whereas gene *ARSA*, located on chromosome 22, remains genome-wide significant in the MA cohort.

**Figure 3.**
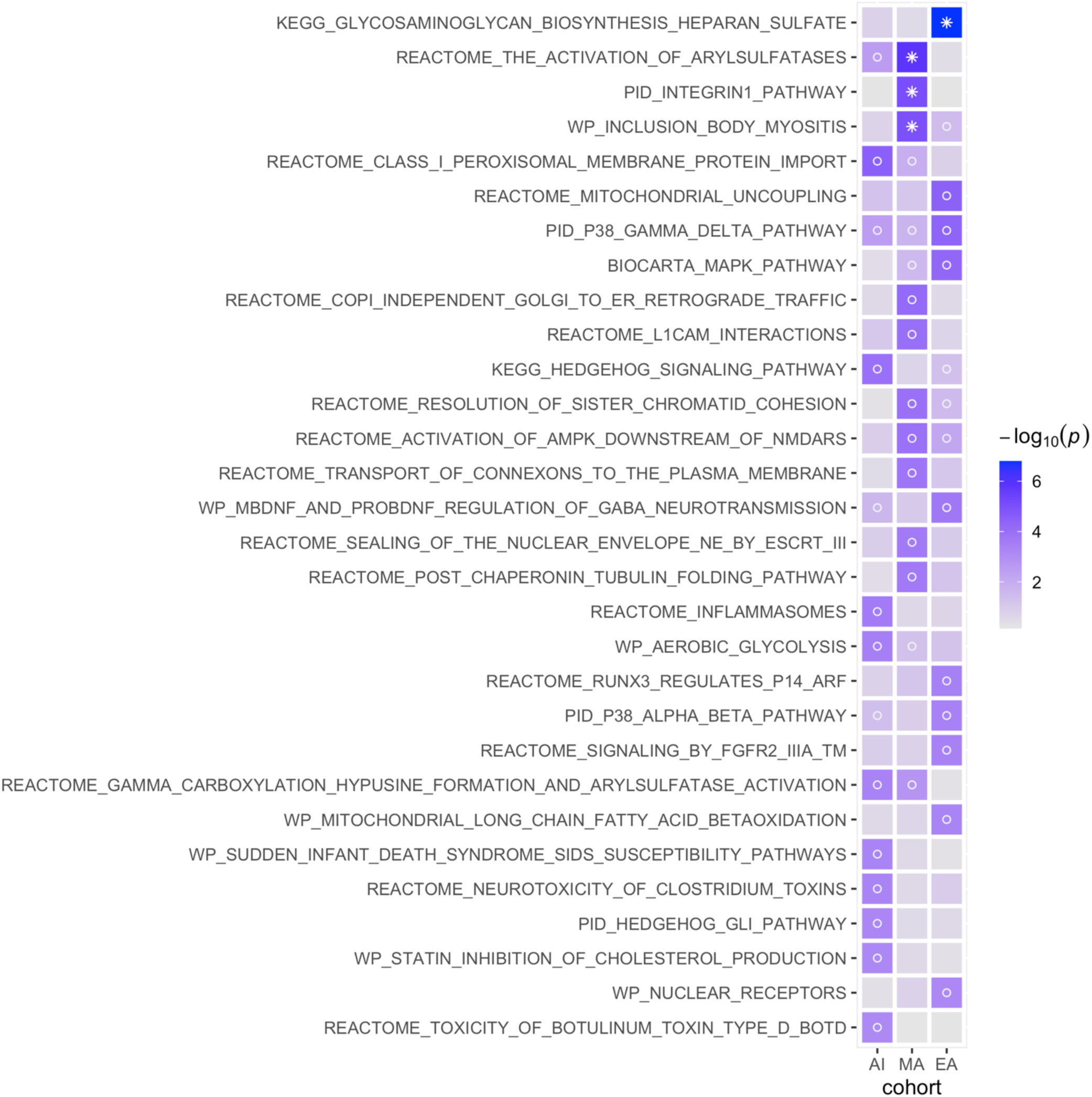
Top pathways enriched for rare and low-frequency variant associations with CUD severity across American Indian (AI), Mexican American (MA), and European American (EA) cohorts. The figure displays the top 10 pathways associated with CUD severity in each cohort. Full details, including the genes within each pathway, are provided in Table S5. Variant selection was limited to 0.1% < MAF < 5%. Cell color indicates the significance level of the association test. Symbols denote significance: * indicates pathways that remain genome-wide significant after FDR correction; o indicates nominal significance.

As illustrated in Figure 4A, seven networks of pathways emerged across the cohorts, formed from the top 10 pathways associated with CUD severity in each of the three cohorts. Pathways within each network cluster share overlapping genes. The size of these networks, defined by the number of distinct pathways, ranged from 1 to 9. Figure 4B illustrates tissue specificity of each network cluster based on differentially expressed genes within the cluster.

**Figure 4.**
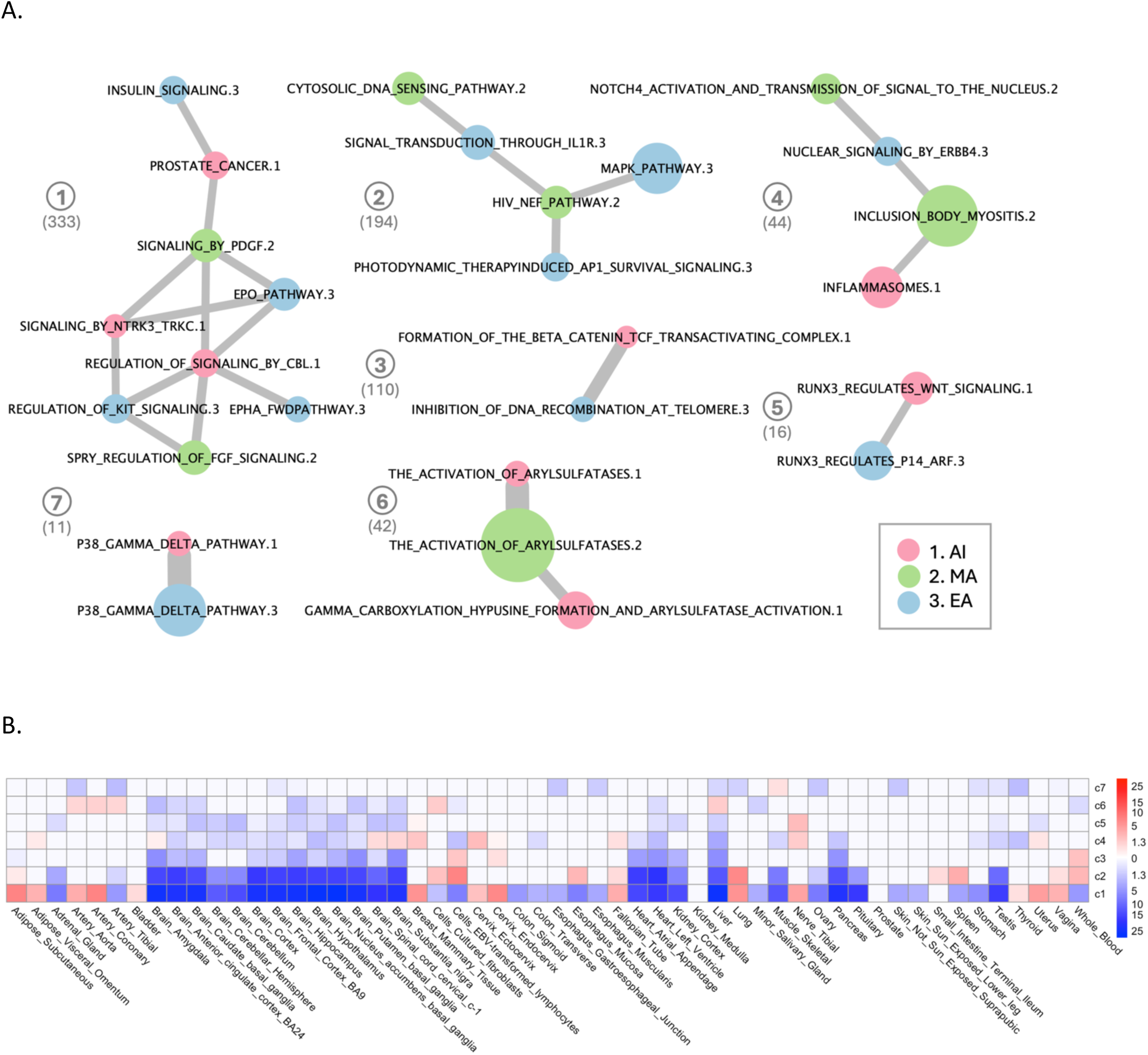
Network clusters of top pathways associated with CUD severity from rare and low-frequency genomic variants across American Indian (AI), Mexican American (MA), and European American (EA) cohorts. A. **Network clusters**: Each circular node represents a pathway, with the database prefixes removed from pathway names for clarity. Pathway names are followed by a number indicating the population cohort (1 = AI, 2 = MA, 3 = EA), which is also reflected in the node color. Node size is proportional to the significance level of the association with CUD severity (-log_10_(p)). Edge width is proportional to the Jaccard coefficient, representing the degree of gene overlap between pathways. The total number of genes in each cluster is shown in parenthesis. B. **Tissue specificity of network clusters**: Each row represents a network cluster from panel A. Colors indicate the significance levels (-log_10_(p)) of differential gene expression in each tissue: red denotes significant upregulation, and blue indicates significant downregulation. Gene expression data are based on tissue-specific enrichment analysis.

## DISCUSSION

Cannabis use and use disorders have been increasing globally in the last two decades [Degenhardt et al., 2016]. This rise is particularly concerning among youth, especially in minority communities like American Indians and Mexican Americans, as early use had been shown to increase the risk of developing use disorders [Ehlers et al., 2007; Roth et al., 2022] and other symptoms, such as depression, in these populations [Gilder and Ehlers, 2012]. The risk for cannabis use disorder is partly genetic, and identifying novel genes and pathways through genome-wide approaches may uncover biological mechanisms related to the disorder, potentially informing targeted interventions.

We analyzed three independent populations—American Indians, Mexican Americans, and European Americans—to identify genetic factors underlying the severity of CUD. The GWAS revealed 10 loci significantly associated with CUD severity in the American Indian cohort, with MAF ranging from 1.3% to 7.1%, but no such associations in the other two cohorts. The rare and low-frequency variant analysis, the first of its kind for a CUD phenotype, identified several significant pathways, some shared across ancestries and others unique to specific populations. The only variant previously identified for CUD in any admixed American population is rs9815757, a rare intergenic variant with a 0.1% MAF [Levey et al., 2023]. This variant is absent in both our AI and MA cohorts.

Among the top variants associated with CUD severity in AI, rs200530573 is located downstream of the gene *NDUFA12*, which encodes a mitochondrial protein (complex 1) involved in the oxidative phosphorylation system in mitochondria. A recent animal study on the effect of tetrahydrocannabinol (THC)—the primary psychoactive cannabinoid in cannabis—on hippocampal neuronal survival and specification found that THC disrupts the expression of mitochondrial proteins. Specifically, several NADH-ubiquinone oxidoreductase subunit proteins were significantly upregulated following THC exposure [Beiersdorf et al., 2020]. Other significant variants are located in or near genes previously associated with alcohol and substance dependence (including cannabis, cocaine, sedatives, stimulants and/or opioids) in African American populations (*SBK3*) [Wetherill et al., 2019], smoking behaviors [Liu et al., 2019] and cortical morphology [Shadrin et al., 2021] in individuals of European ancestry (*PCCA*), brain volume and executive function in Europeans (*FBXO31*) [Hatoum et al., 2023; Zhao et al., 2019], immune system-related blood cell measurements in Europeans and other populations (*ANK1*) [Astle et al., 2016; Chen et al., 2020], and various lipid levels across multiple populations (*SUMO1P1*) [Graham et al., 2021; Sakaue et al., 2021].

### Arylsulfatases activation and heparan sulfate degradation linked to CUD

The activation of arylsulfatases was the most significant pathway, with its functional rare and low-frequency variants associated with CUD severity in the Mexican American cohort. The same pathway, along with another related to arylsulfatase activation, also ranked among the top pathways in the American Indians, where it was nominally associated with CUD severity. The pathway includes genes encoding various arylsulfatases (*ARSA, ARSB, ARSD, ARSF, ARSG, ARSH, ARSI, ARSJ, ARSK, ARSL*), sulfatase-modifying factors (*SUMF1,SUMF2*), and steroid sulfatase (*STS*). Genes in this pathway are significantly downregulated in the hippocampus and amygdala (Figure S5A). Additionally, gene *ARSA* was significant in the gene-based test for the MA cohort. Arylsulfatase A (ARSA) catalyzes the breakdown of sulfatides within lysosomes. Sulfatides are a subgroup of sphingolipids, and play a vital role in white matter lipid homeostasis, crucial for maintaining myelin integrity [Suzuki and Vanier, 1999]. In recent years, sphingolipid systems have emerged as potential pathways involved in addiction development and its pathophysiological consequences [Kalinichenko et al., 2018]. Specifically, sulfatides have been implicated in neurodegenerative process induced by alcohol consumption and nicotine dependence [Yalcin et al., 2017; Yalcin et al., 2015]. Several other arylsulfatases contribute to the degradation of glycosaminoglycans (GAGs), which play key roles in cell signaling [Raman et al., 2005]. For instance, arylsulfatase B (ARSB) breaks down the GAGs dermatan sulfate (DS) and chondroitin sulfate (CS), while arylsulfatase G (ARSG) is essential for heparan sulfate (HS) degradation. Inactivation of ARSG leads to the accumulation of HS in visceral organs and the central nervous system [Kowalewski et al., 2012].

Interestingly, a pathway related to the synthesis of heparan sulfate, a type of GAG, is significantly negatively associated with CUD severity in European Americans, suggesting that more severe CUD may be linked to HS degradation. This pathway was identified in the burden test, where only variants with the same direction of effects were considered. Genes in this pathway are significantly upregulated in fibroblasts and brain tissues, including the frontal cortex, anterior cingulate cortex, and hippocampus (Figure S5B). HS is a component of the cell surface and extracellular matrix (ECM). Evidence suggest that abnormalities in HS biosynthesis can disrupt neuroplasticity, reward signaling, and receptor dynamics, all of which are crucial in the development and persistence of drug-seeking behaviors. Specifically, stimulants like cocaine and methamphetamine have been shown to significantly increase HS abundance and sulfation levels in the lateral hypothalamus, a brain area involved in motivation for both natural rewards and drugs of abuse. HS, in turn, contributes to regulating cocaine seeking and taking behavior. It was demonstrated that HS degradation in the lateral hypothalamus accelerated the acquisition of cocaine self-administration behavior in mice [Chen et al., 2017]. A recent study further showed that stimulants induced significant changes in HS and CS in mouse striatum and lateral hypothalamus, and downregulation of ARSB prevented cocaine preference during withdrawal in mice [Sethi et al., 2024]. Additionally, a study on astrocytes found that ethanol inhibited the ARSB activity [Zhang et al., 2014].

### Integrin cell surface interactions uniquely associated with CUD in Mexican Americans

The integrin β-1 (*ITGB1*, also known as *CD29*) cell surface interaction pathway is significantly enriched in the MA cohort. Integrins are membrane receptors involved in cell adhesion, linking the actin cytoskeleton to the extracellular matrix (ECM) and transmitting signals between the ECM and the cytoplasm [Schwartz et al., 1995]. They are known to play prominent roles in both functional and structural neuroplasticity [DeMali et al., 2003]. Additionally, studies suggest that integrins mediate cannabinoid receptors (CB1)-focal adhesion kinase (FAK) signaling in neuronal cells [Dalton et al., 2020]. As cell surface receptors, integrins are crucial to the pathophysiology of various brain diseases and are considered potential drug targets for neurological disorders [Wu and Reddy, 2012]. The negative association between rare and low-frequency nonsynonymous and splicing site variants within the integrin pathway and CUD severity suggests that these variants may alter integrin signaling, impairing neurons’ ability to adapt to cannabis effects. This disruption could lead to reduction in the severity of CUD, potentially offering new targets for therapeutic strategies.

### Immune response pathways related to CUD with multi-ancestry supports

Inclusion-body myositis (IBM) is an autoimmune muscle disorder characterized by inflammation. This pathway is negatively associated with CUD severity in the MA cohort, which aligns with the immunomodulatory effects of the cannabinoid system, which generally suppresses immune responses [Cabral, 1999; Nichols and Kaplan, 2019]. The pathway also shows nominal significance in the EA cohort. The IBM pathway contains genes linked to inflammation, neuroimmune activity, synaptic plasticity, and the production of amyloid-beta and tau proteins (Table S5). Another noteworthy pathway is the signaling pathway mediated by p38-gamma and p38-delta (Figure 3). This pathway shows suggestive significance in the EA cohort (Table S5) and nominal significance in both the AI and MA cohorts. P38-gamma and p38-delta play key roles in regulating immune cell functions and are essential components of the innate immune response. The associated signaling pathways are crucial regulators of the immune system [Risco et al., 2012].

In addition, seven network clusters of pathways emerged across the three population cohorts when the top pathways associated with CUD in each population were connected based on their shared genes. Genes in most of these network clusters are downregulated in various brain tissues. Furthermore, nearly all clusters show downregulation in the ventricle of the heart and in liver tissues. The liver plays a central role in metabolizing cannabinoids, particularly the high lipid-soluble THC.

### Strengths and limitations

The findings of this study should be interpreted considering both the study design and its limitations. First, the sample sizes are moderate, which limits our statistical power for detection. However, we did identify genome-wide significant variants in the AI cohort, partly due to the higher prevalence of CUD and severe disorders in that population, and partly due to the use of a quantitative trait, which typically provides increased power. Second, while both the AI and EA cohorts had whole genome sequencing data, the MA cohort’s genotype data was derived from an exome array. As a result, the density of rare and low frequency variants in genes is higher in the AI and EA cohorts than in the MA cohort, which may have impacted performance. Despite these limitations, our findings highlight convergent pathways for CUD severity across ancestries. By focusing on the functional mutations within biological pathways, the results remain highly interpretable.

In conclusion, we conducted first large-scale analysis of rare and low-frequency variants in multi-ancestry populations and identified several novel pathways for CUD severity. Our findings indicate that while each population has unique variants and pathways linked to CUD, there are also shared genetic factors across populations. Notably, the rare and low-frequency functional mutations in the activation of arylsulfatases and heparan sulfate degradation cascade may point to an important mechanism underlying CUD development, offering potential therapeutic targets for treatment and intervention.

## DATA AND CODE AVAILABILITY

The EA dataset can be accessed through dbGaP (phs001458.v1.p1). The AI and MA dataset are available by contacting the last author. However, data availability of AI is subject to approval of the specific AI tribes participating.

Canonical pathways GSEA/MSigDB version 7.5.1: http://www.gsea-msigdb.org/gsea/msigdb/download_file.jsp?filePath=/msigdb/release/7.5.1/c2.cp.v7.5.1.symbols.gmt

Hg19 gene list: https://yanglab.westlake.edu.cn/software/gcta/res/glist-hg19.txt GCTA-MLMA version 1.93.2beta: https://yanglab.westlake.edu.cn/software/gcta EPACTS version 3.4.2: https://github.com/statgen/EPACTS.

## Data Availability

The data from the European American cohort under study is available in dbGaP, study accession phs001458.v1.p1. The data from the American Indian cohort under study cannot be shared publicly in accordance with the wishes of the tribes. All other data produced in the present study are available upon reasonable request to the authors.

## ACKNOWLEDGEMENTS

We would like to acknowledge and thank all our research participants, and the following people for their roles in 1) the genotyping effort: Scott Chasse, Piotr Mieczkowski, Ewa Patrycja Malc, Joshua Sailsbery, Phil Owens, and Chris Bizon; and 2) recruiting participants, and collection and preparation of the clinical data: David Gilder, Corinne Kim, Evie Phillips, Gina Stouffer, Susan Lopez, Linda Corey, Greta Berg, Phillip Lau, and Derek Wills.

## AUTHOR CONTRIBUTIONS

QP: conceptualization, methodology, software, formal analyses, validation, writing - original draft, review & editing, visualization, funding acquisition. XZ: data analysis, writing - draft. KW: data acquisition, writing - review & editing, funding acquisition. CE: data acquisition, writing - original draft, review & editing, funding acquisition.

## FUNDING

This work was supported by the National Institutes of Health (NIH): National Institute on Drug Abuse (NIDA) DP1 DA054373 to QP; R01 DA030976 to KCW and CLE. National Institute on Alcohol Abuse and Alcoholism (NIAAA) R01 AA027316 and R01AA026248 to CLE. NIAAA and NIDA had no further role in the study design; in the collection, analysis, and interpretation of data; in the writing of the report; or in the decision to submit the article for publication.

## COMPETING INTERESTS

The authors have nothing to disclose.

## Notes

### Competing Interest Statement

The authors have declared no competing interest.

### Author Declarations

Institutional Review Board of The Scripps Research Institute (TSRI-IRB) gave ethical approval for this work; IRB of the University of North Carolina, at Chapel Hill gave ethical approval for this work.

### Summary of Updates

Main and SI text have been updated.

